# The basic reproduction number and prediction of the epidemic size of the novel coronavirus (COVID-19) in Shahroud, Iran

**DOI:** 10.1101/2020.04.04.20052308

**Authors:** Ahmad Khosravi, Reza Chaman, Marzieh Rohani-Rasaf, Fariba Zare, Shiva Mehravaran, Mohammad Hassan Emamian

**Affiliations:** Ophthalmic Epidemiology Research Center, Shahroud University of Medical Sciences, Shahroud, Iran; Department of Epidemiology, School of Public Health, Shahroud University of Medical Sciences, Shahroud, Iran; Center for Health Related Social and Behavioral Sciences Research, Shahroud University of Medical Sciences, Shahroud, Iran; ASCEND Center for Biomedical Research, Morgan State University, Baltimore, USA

**Keywords:** Epidemic, Reproduction number, Projection, Incident case, COVID-19, Iran

## Abstract

**Objectives:** To estimate the basic reproduction number (R_0_) of COVID-19 in the early stage of the epidemic and predict the expected number of new cases in Shahroud, Northeast of Iran.

**Methods:** The R_0_ of COVID-19 was estimated using the serial interval distribution and the number of incidence cases. The serial interval was fit with a gamma distribution. The probable incidence and cumulative incidence in the next 30 days were predicted using the assumption that daily incidence follows a Poisson distribution determined by daily infectiousness. Data analysis was done using “earlyR” and “projections” packages in R software.

**Results:** The maximum-likelihood value of R_0_ was 2.7 (95% confidence interval (CI): 2.1 to 3.4) for the COVID-19 epidemic in the early 14 days and decreased to 1.13 (95% CI: 1.03 to 1.25) by the end of the day 41. The expected average number of new cases in Shahroud is 9.0±3.8 case/day, which means an estimated total of 271 (95% CI: 178-383) new cases in the next 30 days.

**Conclusions:** It is essential to reduce the R_0_ to values below one. Therefore, we strongly recommend enforcing and continuing the current preventive measures, restricting travel, and providing screening tests for a larger proportion of the population.

## Introduction

At the time of writing this manuscript, the novel coronavirus and the COVID-19 pandemic has already spread to 207 countries worldwide [1, 2], and the number of infected cases continues to escalate. The Islamic Republic of Iran was the first Middle East country to report a case of death due to coronavirus (19 February 2020) and is currently among countries with the highest prevalence of COVID-19. By 3 April 2020, there were 53,183 confirmed cases in Iran, 3,294 of which had already deceased [3]. Given the rapid spread of the virus, the government immediately responded by establishing more than 40 laboratories to enhance the testing capacity, and consequently, there was a sudden spike in the reported number of positive cases. The first cases were immediately reported to the Health Department, and preventive protocols were developed and put in place to limit the further spread of the infection. These included cancelling in-person classes in schools and universities as of 25 February 2020, and switching to online platforms, as well as public awareness campaigns that encourage citizens to minimize face-to-face contact and promote social distancing. Nonetheless, the timeframe from 20 March to 2 April 2020 coincides with the ‘Norouz Spring Holidays’ in Iran. During this time, there is significantly higher rates of social activities, visiting family and friends, trips, shopping, and festivals. This is while the epidemic has already spread throughout the country and beyond. Therefore, close monitoring and evaluation is necessary to investigate the efficiency of control measures, determine the potential community transmission patterns, and predict the progression of the epidemic and the trajectory of the epidemic curve.

One useful epidemic measure which can help investigate the transmissibility of infection is the reproduction number. The basic reproduction number (R_0_) is the average expected number of new cases infected by a primary case and must be estimated early during an epidemic [4]. R_0_, can be affected by various factors such as the probability of transmission upon contact between an infected case and a susceptible person, the frequency of contact, the duration of infection in a person, and the proportion of immune people in the population [5]. The serial interval (SI) of an infection is the mean duration between symptom onset of two successive cases (the primary case and secondary case). The force of infection (denoted λ), which describes the rate at which susceptible people acquire a given infection, is another useful parameter when implementing preventative measures [6].

According to the latest report of the Ministry of Health and Medical Education in Iran, the incidence rate of COVID-19 has been highest in Semnan Province (118 cases per 100000 persons) the highest incidence rate in Iran by April 01 was seen in Shahroud County [7]. Shahroud, in Shahroud County and Semnan Province, is a city located in the northeast of Iran with a population of about 218628 in 2016 [8]. The first confirmed case of COVID-19 in Iran was identified on February 19 in Qom which is about 550 km from Shahroud (Figure 1) [9]. Four days later (February 23, 2020), nasopharyngeal and throat swabs of five suspected cases in Shahroud were submitted for viral nucleic acid testing, and two tested were positive. One of these primary cases was a 74-year-old woman who had been hospitalized on February 10, with chief complaints of fever and cough, and a travel history to Qom. This indicates that the epidemic probably started almost one month before it was known to the public. Given the high incidence rate of COVID-19 in the region, the aim of this report is to estimate the R_0_ of the COVID-19 in the early stage of the epidemic and predict the trajectory of the epidemic and new cases in Shahroud.

**Figure 1:**
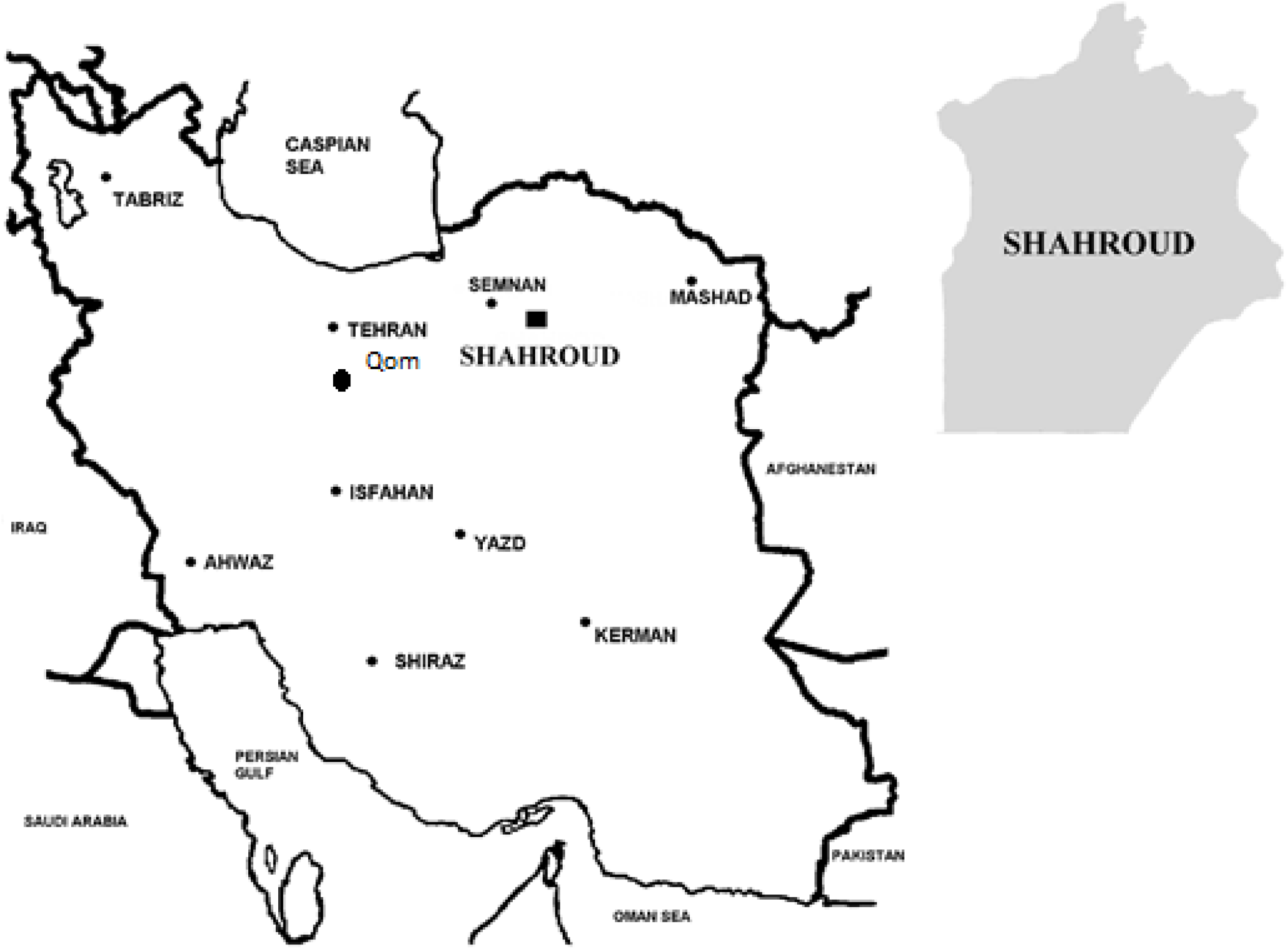
Map of Iran showing the locations of Shahroud in Semnan Province and Qom, where the first case of COVID-19 was identified (road distance=550 km).

## Methods

The protocol of this study was reviewed and approved by the Institutional Review Board of Shahroud University of Medical Science (IR.SHMU.REC.1398.160). The study was conducted at Imam Hossein hospital which is currently the only specialty hospital designated to COVID-19 patients in Shahroud. In the past month, all walk-in and referral cases have first been screened. Of these, suspected cases have been admitted and tested for the infection testing. For testing, two respiratory tract samples (throat and nasopharyngeal swabs) are collected and submitted for viral nucleic acid testing. All positive cases are systematically recorded in a designated registry which is used for follow-up and contact tracing.

In this study, we used an informative prior distribution for the SI, which was estimated as 7.5 ±3.4 days for COVID-19 in Wuhan, China [10], fit with a gamma distribution.

We calculated the likelihood-based R_0_ using a branching process with Poisson likelihood. Bootstrapping with 1000 times resampling was used for obtaining the distribution and confidence interval (CI) of R_0._

We then used the estimates of R_0,_ SI, and daily incidence to simulate the trajectories and project the future daily cumulative incidence where the main assumption was that the model follows a Poisson distribution [11]. For each date *t* ≥ 2, the number of incident cases *I*_*t*_ was drawn from a Poisson distribution with mean 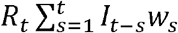, where *R*_*t*_ is the instantaneous reproduction number, *w*_*s*_ is the discrete SI distribution and *I*_*t-s*_ is the incidence at time step *t-s*.

For a 30-day projection, we used a uniform distribution of 0.8 to 1.5 for R_0_ and Bootstrapping with 1000 times resampling. [11, 12] Data analysis was performed using the “incidence”, “earlyR”, “ggplot2” and “projections” packages in R (3.6.3) software.

## Results

During the first 41 days of the epidemic (20 February to 31 March 2020), a total of 1055 suspected samples were tested for COVID-19 in Shahroud, and 405 (40.1 percent) of them tested positive. The daily distribution of these confirmed cases is illustrated in Figure 2.

**Figure 2:**
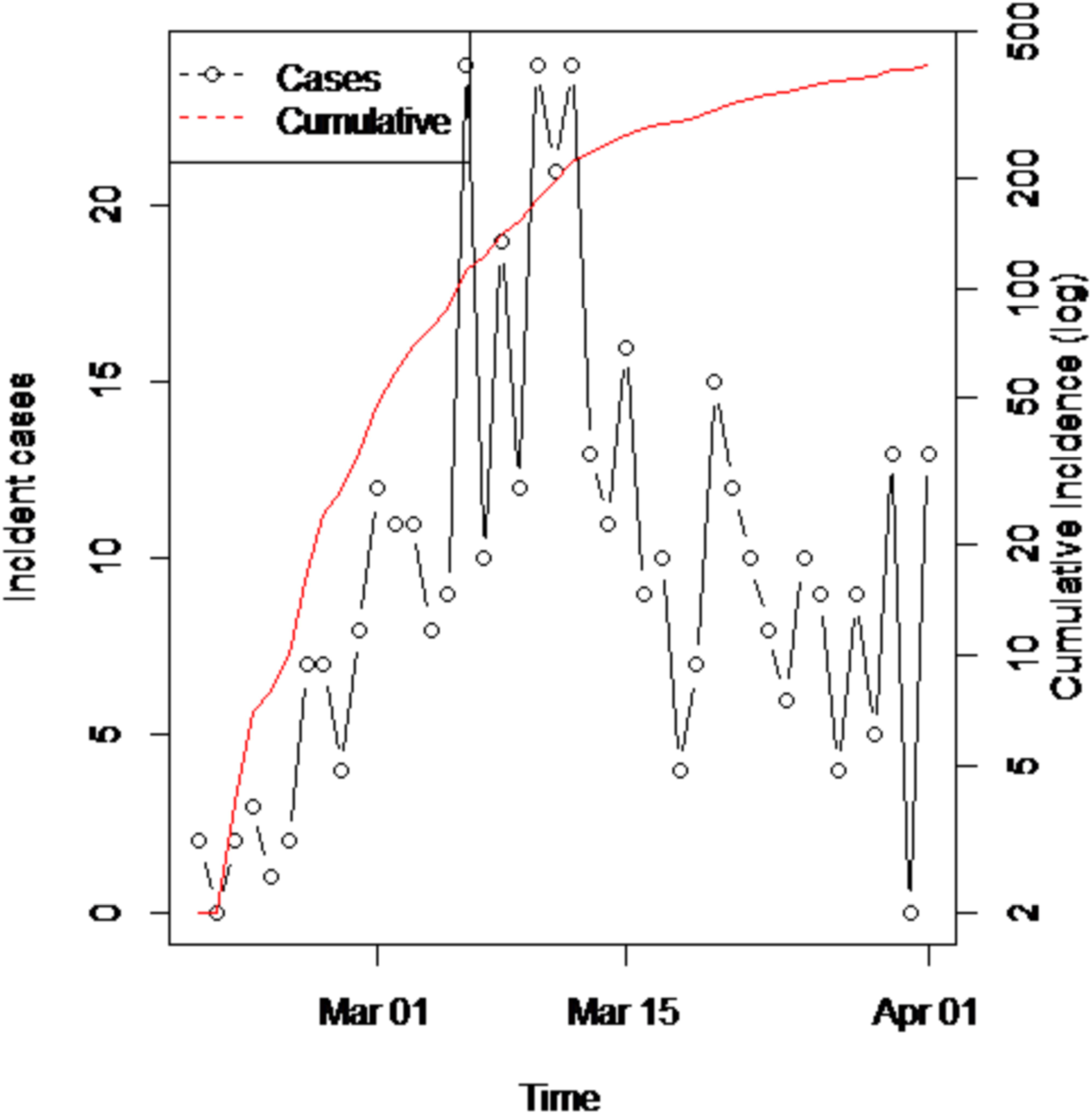
Incidence and cumulative incidence of Covid-19 between 20 February and 31 March 2020 in Shahroud, Iran

Using the SI distribution, the maximum likelihood value of R_0_ was estimated at 2.7 (95% CI, 2.1 to 3.4) which is indicative of a propagated epidemic (Figure 3). To stabilize this estimate, we used a 2-week time window, and the maximum likelihood value of R_0_ decreased to 1.28 (95% CI: 1.14 to 1.43) for day 30 (20 March 2020) and 1.13 (95% CI: 1.03 to 1.25) for day 41 (31 March 2020).

**Figure 3:**
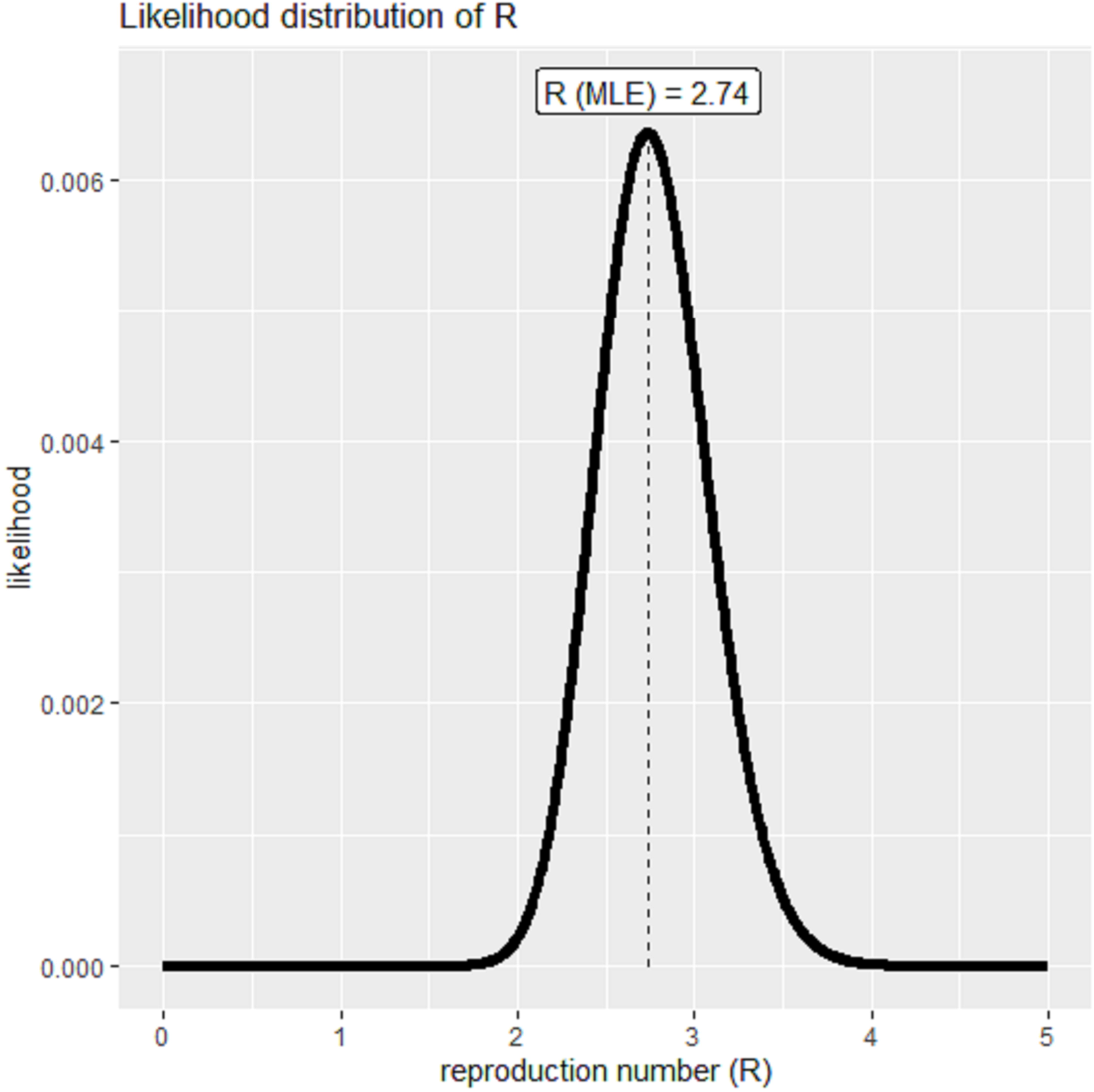
The distribution of likely values of basic reproduction number (R_0_) with the maximum-likelihood estimation.

**Figure 4:**
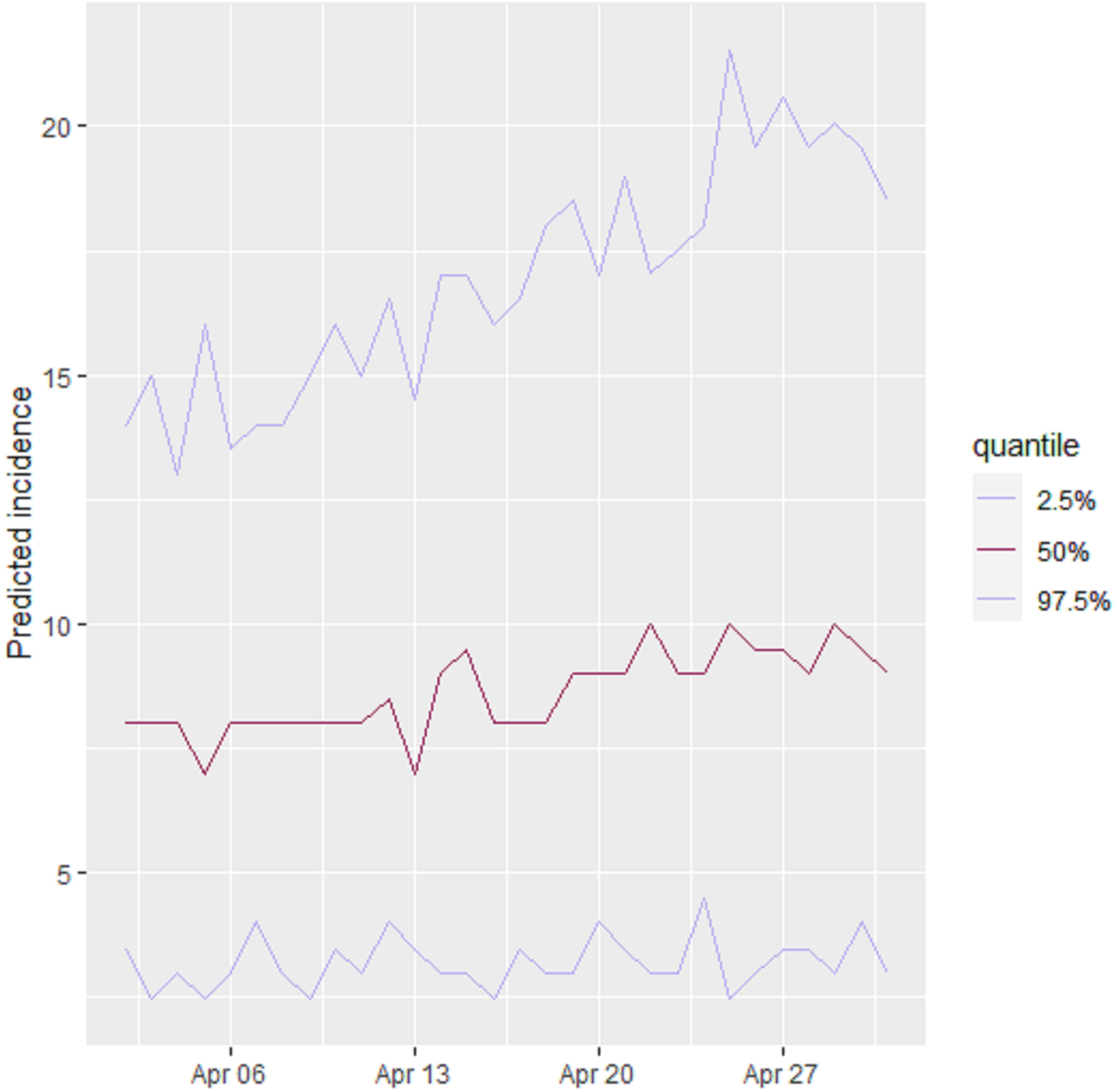
Thirty-day projections of the incidence and cumulative incidence of COVID-19 in Shahroud, Iran. A) Predicted number of incident cases, B) Smoothed number of predicted cases, C) Cumulative incidence if the basic reproduction number follows a uniform distribution of 0.8 to 1.3.

The predicted number of new cases for the next 30 days (one month after ending the Norouz Spring Holidays), based on decreasing function of R_0_ between 1.3 and 0.8, is demonstrated in figure 5 (A, B, C). The overall predicted average number of new cases will be 9.0±3.8 cases per day in the next 30 days. In figure 5-B, the daily average of predicted incident cases was smoothed for the time span. The next 30-day projected cumulative incidence in Shahroud is shown in figure 5-C; approximately 271 (95% CI: 178-383) new cases will be infected in the next 30 days.

## Discussion

The R_0_ of an infection is commonly used to characterize its transmissibility during an epidemic. The trend of R_0_ over time provides a measure of the effectiveness of control and prevention strategies in the community, and to control an epidemic, the goal is to reduce and keep R_0_ below the value of 1 [13]. In the present study, the estimated R_0_ of 2.7 (95% CI, 2.1 to 3.4) during the early stage of the epidemic is in line with previous estimates [10, 14-16]. However, higher estimates of R_0_ have been reported in larger populations [17, 18]. For precise estimation of R_0_, certain conditions must be met which include the precise detection of cases in the early stages of the epidemic, restricting the calculation to a small timeframe [12], and choosing the appropriate estimation method [13, 19]. For precise detection of cases, all suspected cases (according to the screening protocol) and cases who have had close contact with confirmed cases should undergo viral nucleic acid testing. In Shahroud, there were 1055 suspected cases, and 405 of them tested positive. However, in the early stages of the epidemic in Iran, there was limited capacity for testing, and the calculated R_0_ may be an underestimation.

The results of this study showed that R_0_ has decreased temporally. This pattern, which is promising for controlling an epidemic, is due to interventions enforced by the health system starting from the early days of the epidemic. Some of the most important measures were public education to promote social distancing and encouraging people to stay home. In addition, two other basic measures were taken in Shahroud: 1) At the time of hospital discharge, all patients and their caregivers were provided with counseling and training on how to be isolated at home for 14 days, and families received information about how to care for patients; 2) Active contact tracing with follow-up of patients’ family members and friends, work colleagues, and other possible contacts and referral of suspected cases to medical centers [9].

According to our 30-day projection, there should be a decrease in R_0_, and in the next 30 days, 271 cases is expected. This can be due to spreading of disease by unidentified asymptomatic cases and increasing the number of tests performed on outpatients following the improvement of laboratory facilities. So, we strongly recommend measures to identify these cases.

This study can inform health policymakers of the success of the preventive measures and interventions. It also emphasizes the need for these measures to be continued along with stricter limitations in transportation until the transmission chain is broken and the epidemic is successfully controlled.

The main strengths of this study include its careful design, taking throat and nasopharyngeal swabs for testing of all suspected cases, and systematic recording of positive cases. Limitation were that testing was only done for those admitted into the hospital, as well as potential limitations in the calculation of R_0_,

In conclusion, the R_0_ of COVID-19 in Shahroud was considerably high at the onset of the epidemic, but with preventive measures and public education, it has been reduced to 1.13 (95% CI: 1.03-1.25) within 41 days. This reduction highlights the success of preventive measures in place, but we must be prepared for the doubling of cases over the next month. We strongly recommend performing mass screening of suspected cases, implementing travel restrictions especially during Spring holidays, and expanding coronavirus testing to the community. After the ending the holidays, longer and stronger limitations needed.

## Data Availability

Data are available on request

## Acknowledgements

This work was supported by Shahroud University of Medical Sciences (Grant No. 98126).

## Conflicting Interests

The Authors declare that there is no conflict of interest.

---

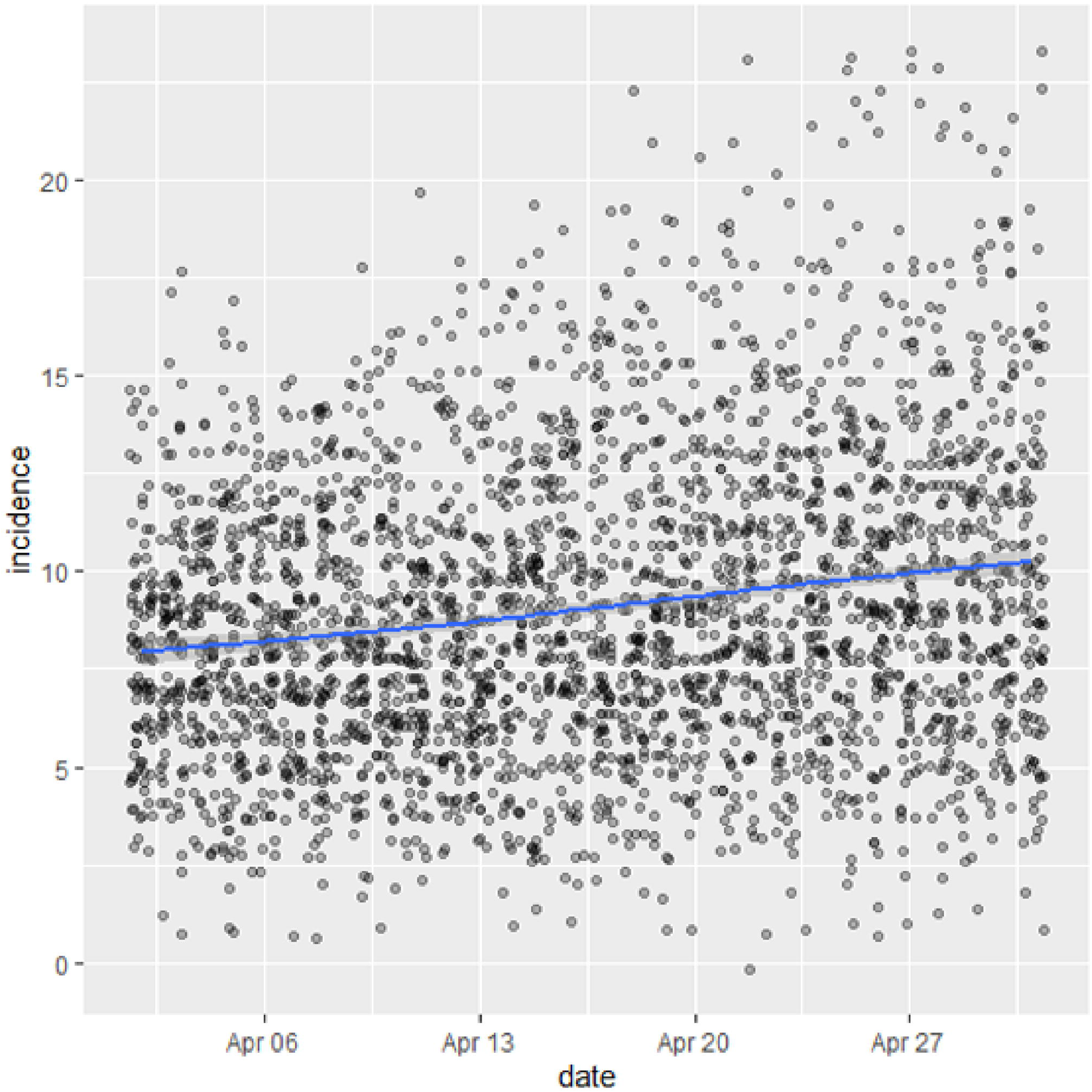

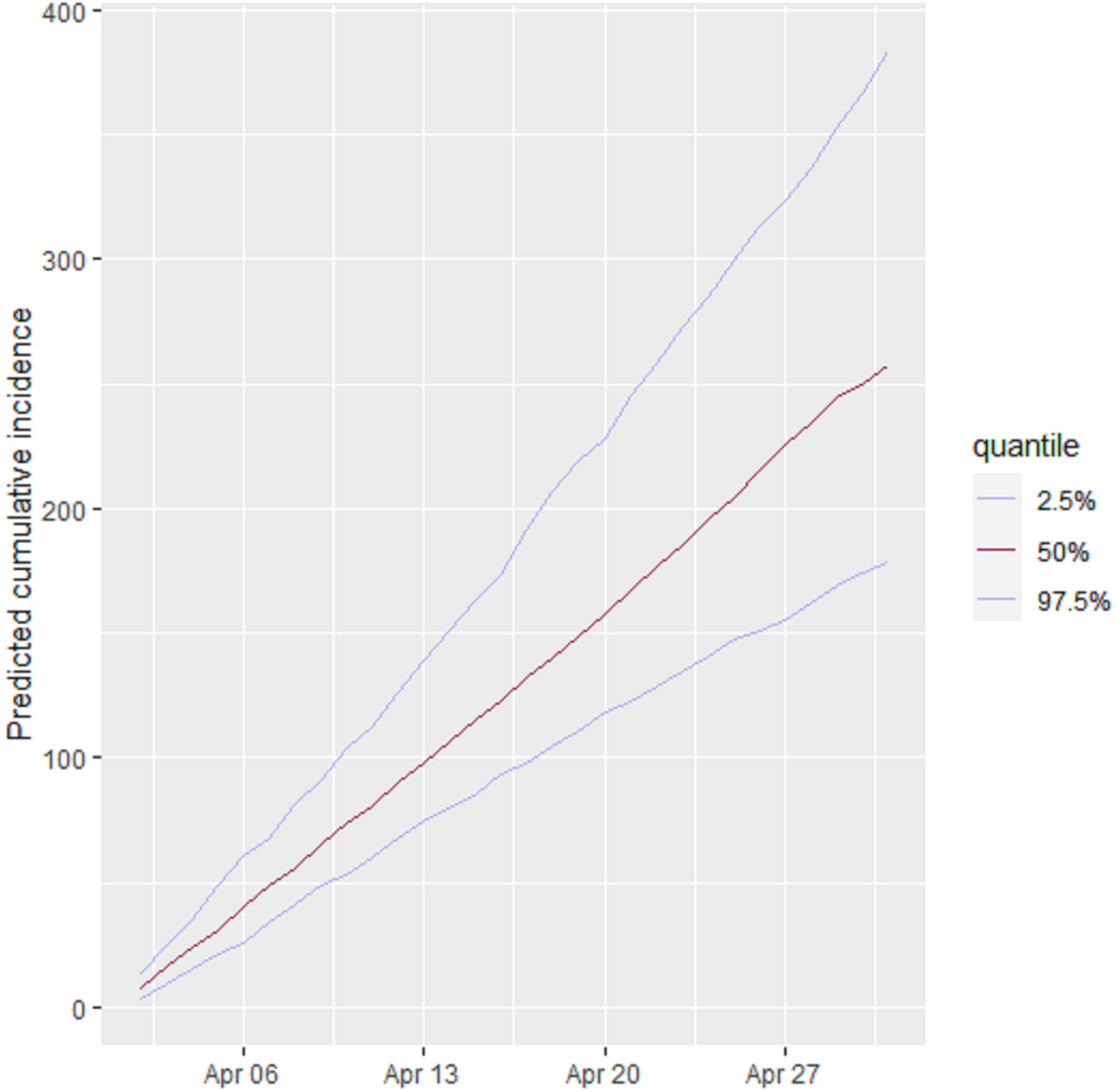

## Notes

### Competing Interest Statement

The authors have declared no competing interest.

